# Triglyceride- glucose index as simple marker associated with atherosclerotic cardiovascular disease in a tertiary care hospital in Pakistan

**DOI:** 10.1101/2024.04.22.24306160

**Authors:** Maliha Akhtar Zubairy, Muhammad Nizamuddin

**Author notes:** **Corresponding Author:** Maliha Akhtar Zubairy, Consultant Chemical Pathologist, Indus Hospital and Health Network.

## Abstract

Identification of patients at early risk for CVD improves risk stratification and patient management. Triglyceride glucose index can be used as a marker of atherosclerotic cardiovascular disease.

This study was approved by IRB of the institute and conducted in the period of July 2020 to June 2022 in the Chemical Pathology laboratory of Indus Hospital. All patients whose cardiac intervention (PCI/LHC) performed were included. Fasting lipid profile was analyzed on Alinity C analyzer, and FBS was measured on glucometer.

TyG index was calculated, 54% have TyG index more than 9.04. Male predominance was observed, with 68.6% falling within the <9.04 range and 60.4% within the >9.04 range. Patients aged over 50, constituting 51 (59.3%) in the <9.04 TG index group and 60 (59.4%) in the >9.04 TG index group.

Patients with TyG index <9.04, majority FBS is in pre-diabetic range 45.3% while TyG index >9.04 group, majority FBS is in diabetic range with a p-value of <0.001.

Our study demonstrated that the TyG index was independently associated with atherosclerosis in our population and this marker can be used for the prediction of disease for healthy individuals.

## Introduction

Cardiovascular disease (CVD) is a leading cause of morbidity and mortality, with serious public health challenges. Multiple risk factors for CVD have been established like advancing age, male sex, obesity, hypertension, hypercholesterolemia, and diabetes mellitus. Research has given evidence that without these risk factors, the chances of cardiovascular disease are still present in the population (1).

Identification of patients at early risk for CVD improves risk stratification and patient management. The risk is associated with insulin resistance and related disorders like hyperglycemia, dyslipidemia, and hypertension (2, 3 and 4). Insulin resistance (IR) is the principal feature of metabolic syndrome and it can independently predict the development of CVD (5, 6, 7, 8). Thus, IR is a pathogenic cause and also a predictor of CVD. Insulin resistance is the main feature and has a significant pathogenic link to type 2 diabetes mellitus (T2DM) (3). Therefore, the early identification of individuals with insulin resistance will be essential to reduce the disease burden of CVD {2} Increasing evidence in postmenopausal women suggests a protective effect of endogenous estrogen against CAD, whereas its deficiency exacerbates the process of CAD after menopause (6).

Therefore a reliable and convenient screening tool is required to detect IR and predict cardiovascular risk. Currently, the gold standard test is the euglycemic insulin clamp and intravenous glucose tolerance test, which are invasive expensive, and not applied in clinical practice (8). The homeostasis model assessment estimated insulin resistance (HOMA-IR) index is widely used but it has limited value in patients with insulin and dysfunctional beta cells.

To address this limitation, the triglyceride glucose (TyG) index has been developed and was shown to be superior to HOMA-IR in assessing IR in individuals with and without diabetes. The triglyceride-glucose (TyG) index, which is the logarithmic product of fasting triglyceride and glucose, is a simple measure of insulin resistance (2) that a higher TyG index was significantly associated with an increased risk of future myocardial infarction (MI), and stroke in a study on 7,183,262 persons aged 40 years and older who participated in the national health screening program. The TyG index has been established in healthy young adults as an effective screening tool for IR (5).

This simple, convenient, and low-cost marker does not require insulin levels and can be used in all subjects despite insulin treatment. Studies have demonstrated that the TyG index is an independent predictor of prognosis in diabetic or nondiabetic patients with CVD, suggesting its potential clinical utility in predicting cardiovascular risk.

### Objectives

a) To classify patients according to the Triglyceride Glucose Index as low and high-risk patients for atherosclerotic cardiovascular disease.

d) To determine the association of high TGI with age, gender, HbA1c, FBS, cholesterol, heart vessel disease, HDL, and LDL.

### Operational definitions

#### 1. Atherosclerotic heart disease

According to the American Heart Association atherosclerotic cardiovascular disease, otherwise known as ASCVD, is caused by plaque buildup in arterial walls and refers to conditions that include: Coronary Heart Disease (CHD), such as myocardial infarction, angina, and coronary artery stenosis.

#### 2. Diabetes Mellitus

According to American Diabetes Association Guidelines 2023: Criteria for the diagnosis of diabetes:

FPG ≥126 mg/dL (7.0 mmol/L). Fasting is defined as no caloric intake for at least 8 h. **OR**
**OR**
A1C ≥6.5% (48 mmol/mol). The test should be performed in a laboratory using a method that is NGSP-certified and standardized. **OR**

#### 3. Triglyceride Glucose Index

The TyG index was calculated as the natural logarithm (Ln) of the product of plasma glucose and TG using the formula: Ln (TG [mg/dL]×glucose [mg/dL]/2).

Reference: Romero FG, Mendia LES, Ortiz MG, Abundis M, Zavala R, Hernandez Gonzalez H, *et al*. The product of triglycerides and glucose is a simple measure of insulin sensitivity. Comparison with the euglycemic–hyperinsulinemic clamp. J Clin Endocrinol Metab, 95 (2010), pp. 3347-3351

#### 4. Hypercholesterolemia

ATP III Classification of Serum Cholesterol (mg/dL)

200-239 Borderline high

<200 Normal

#### 5. Hypertriglyceridemia

ATP III Classification of Serum Triglycerides (mg/dL)

<150 normal

150-199 Borderline High

200-499 High

=>500 Very High

#### 6. Hyperlipidemia

Fredrickson DS. An international classification of hyperlipidemias and hyperlipoproteinemias. Ann Intern Med. 1971 Sep;75(3):471-2:

“Hyperlipidemia as low-density lipoprotein (LDL), total cholesterol, triglyceride levels, or lipoprotein levels greater than the 90th percentile in comparison to the general population, or an HDL level less than the 10th percentile when compared to the general population. Lipids typically include cholesterol levels, lipoproteins, chylomicrons, VLDL, LDL, apolipoproteins, and HDL.”

### Duration of data extraction

June 2020 to June 2022

### Setting

Chemical Pathology laboratory, The Indus Hospital.

### Study design

Retrospective chart review.

### Rationale

Through this study, we will determine that atherosclerotic cardiovascular diseases are associated with Triglyceride glucose index (TyG index) in inpatients of The Indus Hospital. We will identify the situation of possible risk of ASCVD in our population to guide the community.

### Inclusion criteria

Age >=18 years and both genders. All the test samples received for FBS and Fasting lipid profile from OPD and IPD

### Exclusion criteria

Age <18 years and pregnant patients.

## Materials and methods

The study data will be extracted from the hospital management information system HMIS in Microsoft Excel format for patient history, FBS, Fasting lipid profile, location, and department from June 2020 to June 2022.

The study was approved by the Institutional Review Board (IRB) of the Indus Hospital Karachi. The retrospective data was collected from electronic medical records. A total of 386 inpatients for which fasting triglycerides and fasting blood sugar were ordered by consulting Physicians were included. For confirmation of atherosclerotic heart disease, only inpatients, whose percutaneous coronary intervention (PCI) and or Left heart catheterization (LHC) performed were included in the study. Triglycerides, LDL, and HDL were analyzed by spectrophotometric assays on an Alinity C analyzer, and fasting blood glucose was measured on a glucometer by biosensor chip technology for Triglyceride Glucose index calculation. For a few patients, for which glucometer blood sugar results were not available, Fasting blood glucose results from the laboratory, were used for triglyceride glucose index calculation (included from data extracted via EMR).

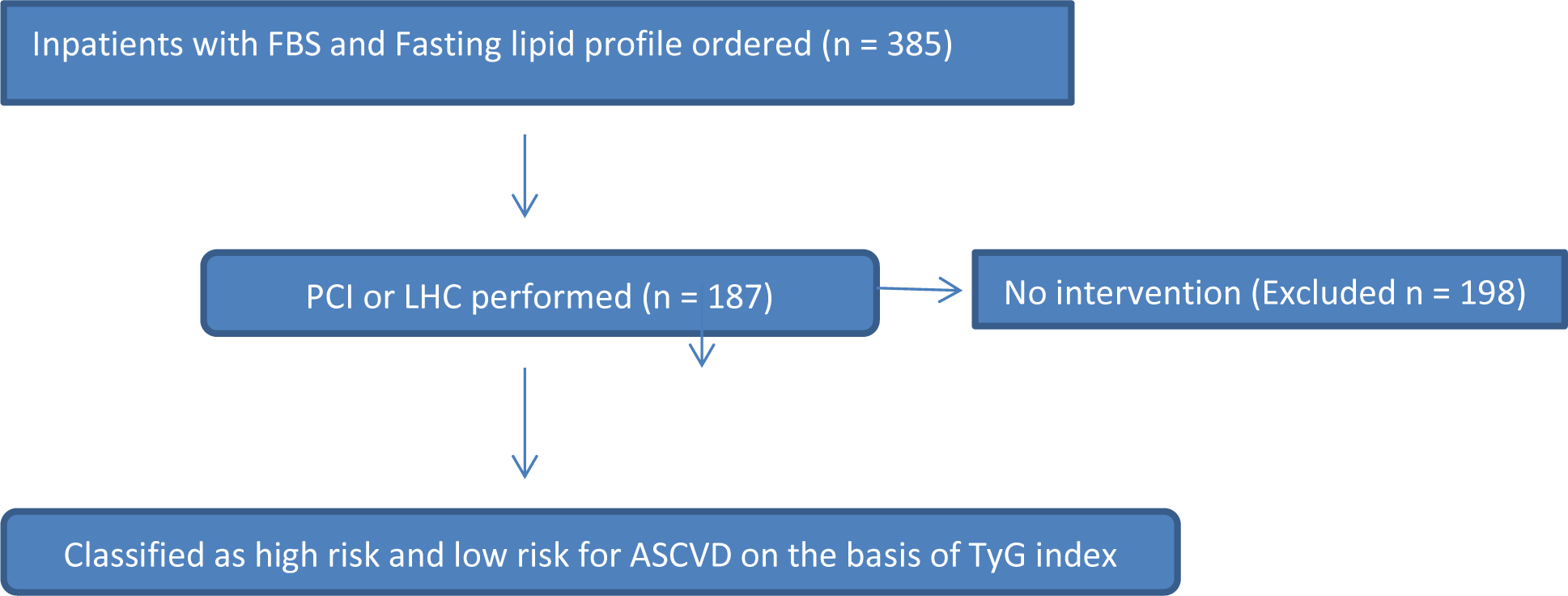

The TyG is calculated by the formula:

TyG index = ln [fasting triglyceride (mg/dL) × fasting glucose (mg/dL)]/2.

## Results

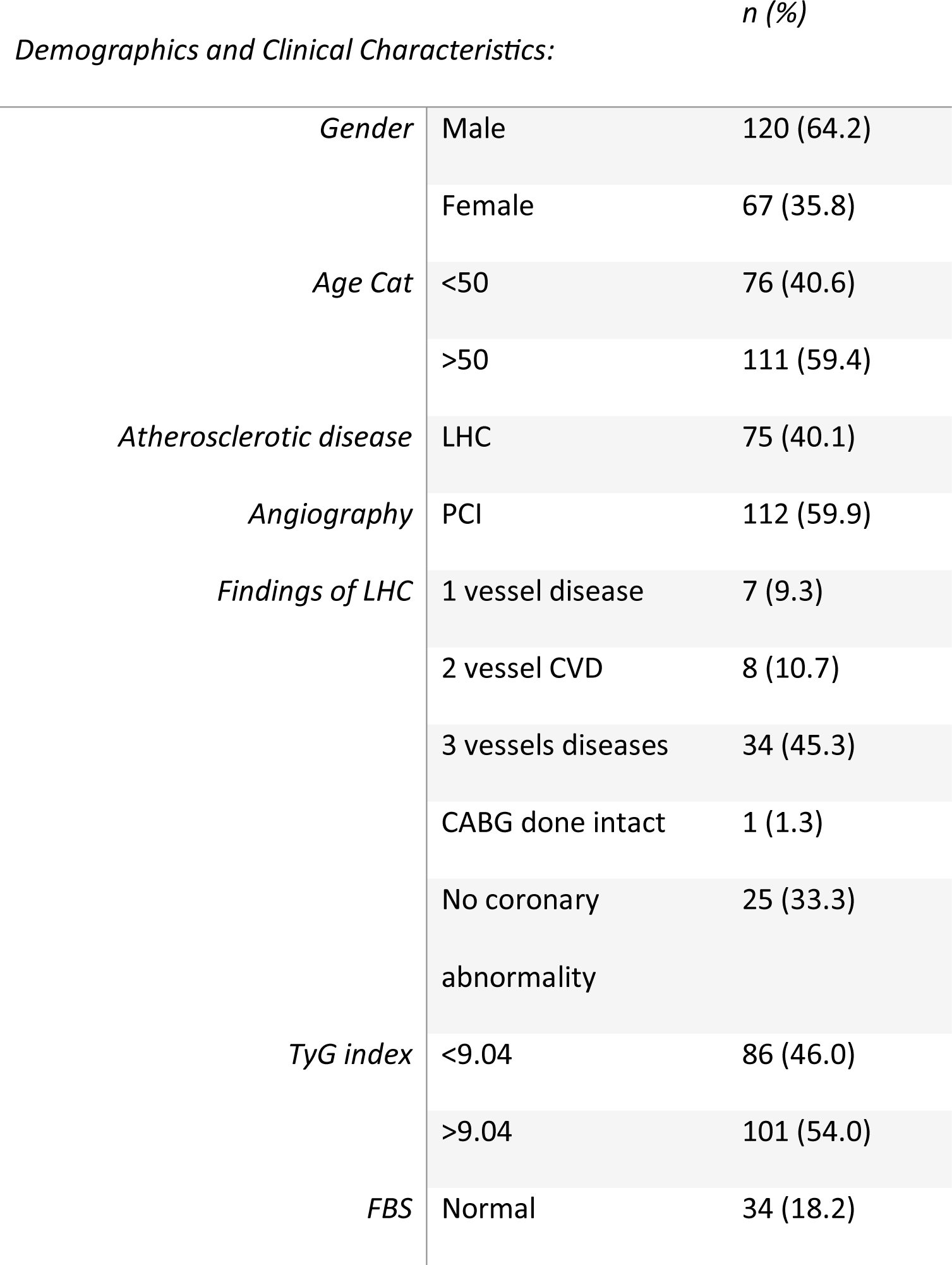
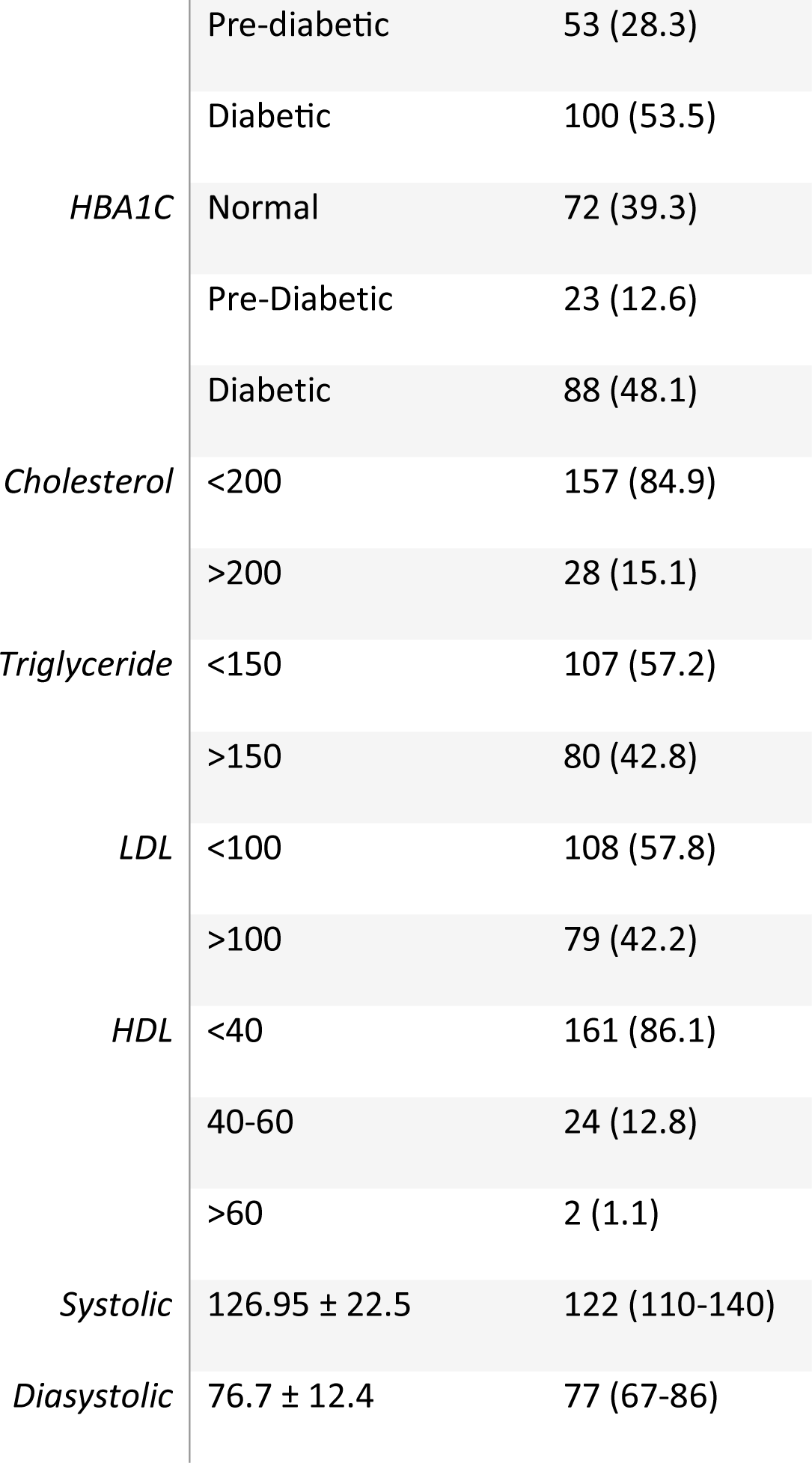

The predominant demographic in the study comprised males, accounting for 120 individuals (64.2%). The age distribution was that the majority was aged higher than 50 years, 111 (59.4%). Percutaneous Coronary Intervention (PCI) was the chosen intervention in 112 cases (59.9%), whereas 75 cases (40.1%) underwent Left Heart Catheterization (LHC). Within the LHC subgroup, 3-vessel disease was notably prevalent, documented in 34 individuals (45.3%). The established cutoff values were delineated as <9.04 to denote normal levels and >9.04 to indicate elevated levels of the triglyceride index. Out of the total patients, a majority of 101 individuals (54%) surpassed the defined threshold, demonstrating a triglyceride index that exceeded the normative value of 9.04.

The laboratory parameters, expressed as frequencies and percentages (%), encompassed the following: Fasting Blood Sugar (FBS) in the diabetic range was observed in 100 individuals (53.5%), while elevated Hemoglobin A1C (HBA1C) levels indicative of diabetes were found in 88 subjects (48.1%). Cholesterol levels below 200 mg/dL were recorded in 157 participants (84.9%). Triglyceride levels were below 150 mg/dL in 107 individuals (57.2%), and High-Density Lipoprotein (HDL) cholesterol was less than 40 mg/dL in 161 subjects (86.1%). Low-density lipoprotein (LDL) cholesterol levels were below 100 mg/dL in 108 participants (57.8%). Additionally, Systolic Blood Pressure ranged from 110 to 140 mmHg with a median of 122 mmHg, while Diastolic Blood Pressure ranged from 67 to 86 mmHg with a median of 77 mmHg, as delineated in Table 1.

In both categories of the TG glucose index, male predominance was observed, with 59 (68.6%) falling within the <9.04 range and 61 (60.4%) within the >9.04 range. A substantial proportion of patients were aged over 50 years, constituting 51 (59.3%) in the <9.04 TG index group and 60 (59.4%) in the >9.04 TG index group.

Among patients in the TyG index <9.04 group, the majority exhibited fasting blood sugar (FBS) levels pr-diabetic, accounting for 39 (45.3%). Conversely, within the TyG index >9.04 group, the majority demonstrated FBS levels diabetic, totaling 76 (75.2%). This disparity yielded statistical significance, with a p-value of <0.001.

Patients with controlled HBA1C were predominantly distributed within the TyG index <9.04 category, encompassing 48 (57%), while in the TyG index >9.04 group, the majority exhibited uncontrolled HBA1C levels, comprising 65 (65.7%). This discrepancy was statistically significant, with a p-value of <0.001.

A minority of patients, specifically 5 (5%), with a TyG index <9.04 exhibited cholesterol levels >200, whereas, within the TyG index >9.04 category, 23 (23%) displayed cholesterol levels >200. This variation reached statistical significance, with a p-value of <0.001.

The majority of patients, encompassing 82 (95%), with a TyG index <9.04 manifested triglyceride levels <150. Conversely, among patients with a TyG index >9.04, 76 (75.2%) demonstrated triglyceride levels >150. This discrepancy was statistically significant, with a p-value of <0.00, table 2.

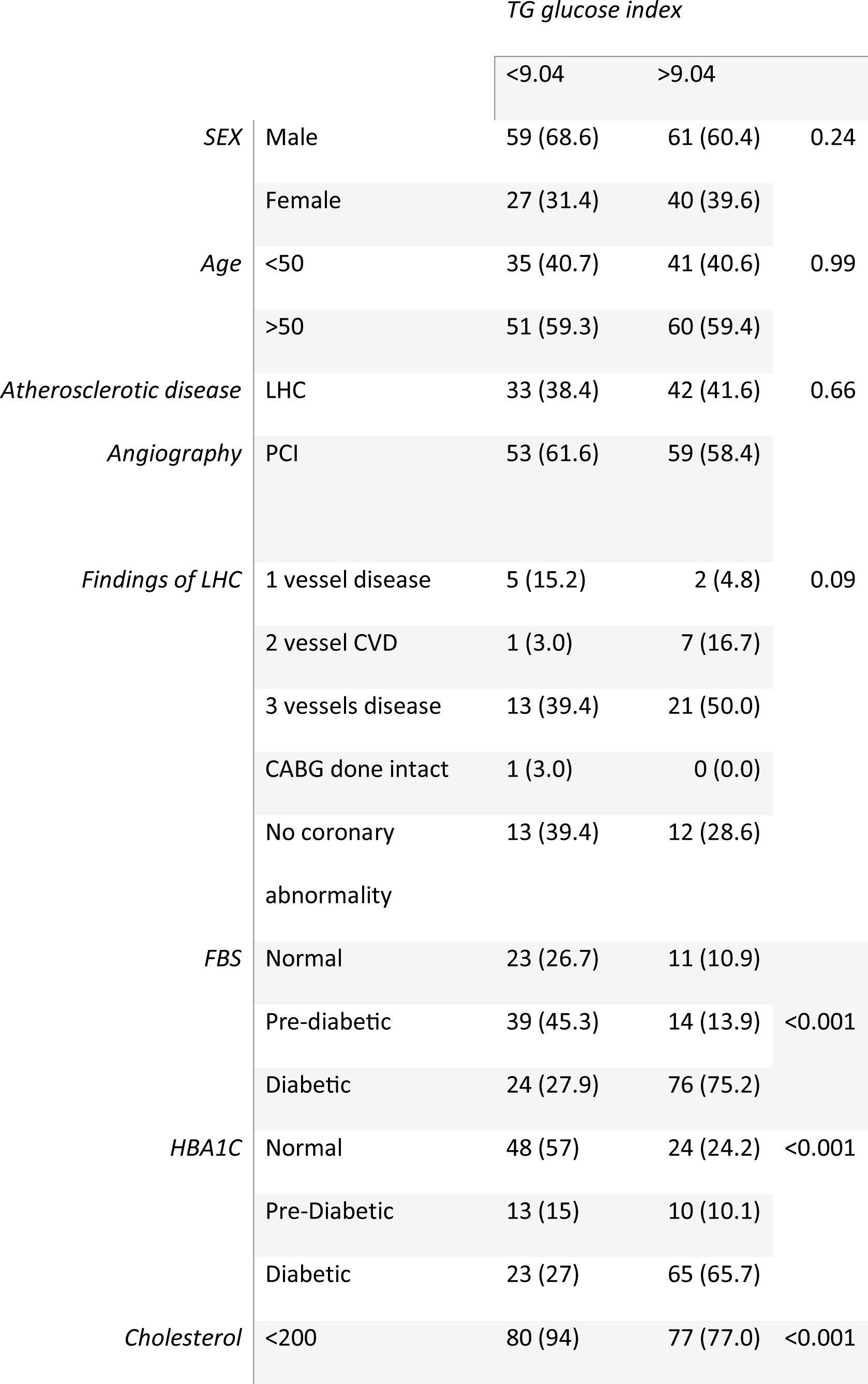
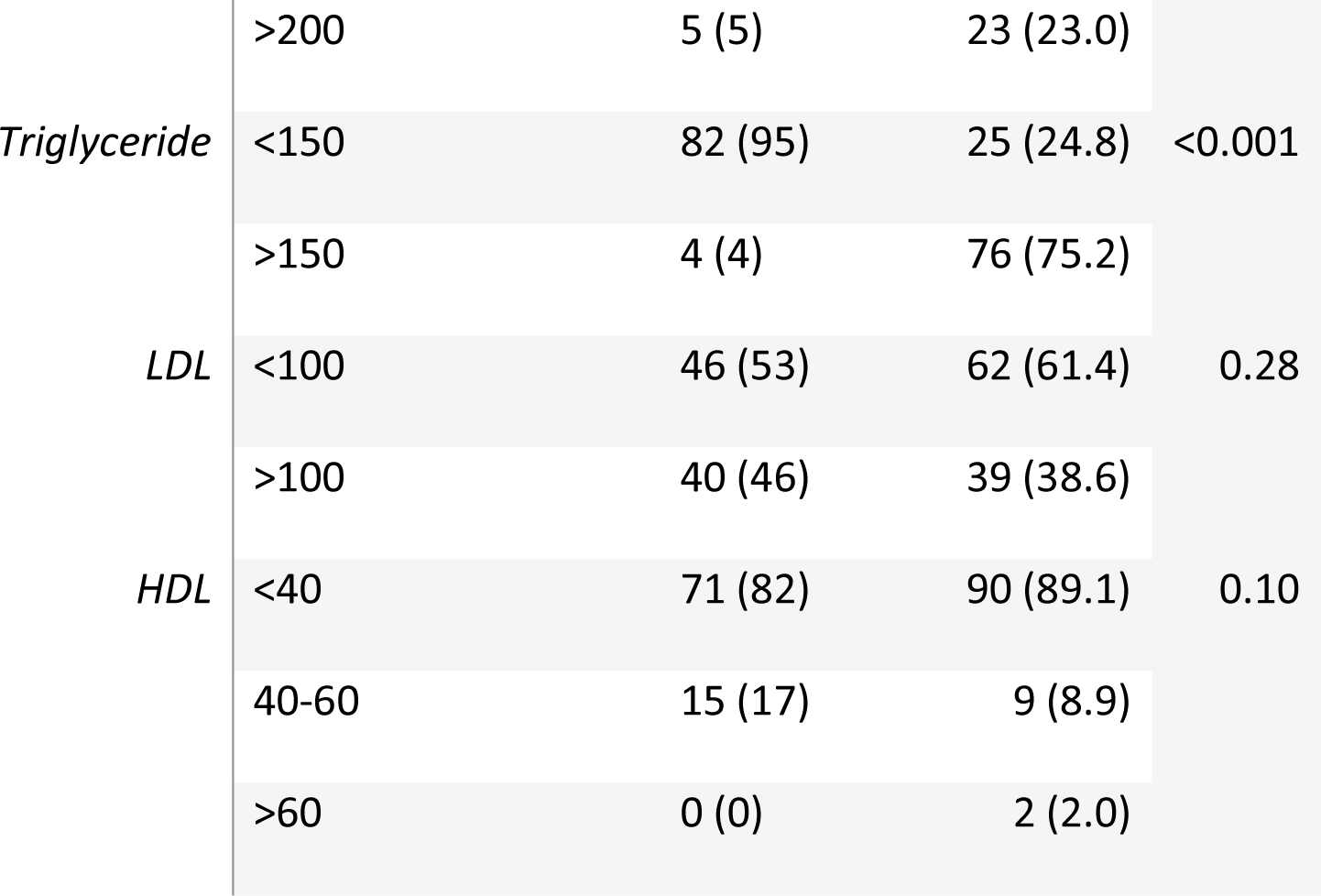

### Statistical Analysis

The results will be analyzed using SPSS. Descriptive statistics will be used for quantitative variables whereas qualitative data will be evaluated by frequency and percentage. The frequency of the high and low TG Index is calculated by the TyG index formula. The data will be calculated as quarters of a year (three months) and compared. The correlation of low and high GI with age and gender will be calculated and the p-value calculated (p-value <0.05 will be considered statistically significant). The association of co-morbid conditions with the TyG Index will be calculated by chi-square analysis or Fisher Exact test. P-value<0.05 will be considered statistically significant.

## Discussion

The study was performed in a tertiary care hospital inpatients. Triglyceride glucose index was used as a simple surrogate marker of insulin resistance in the population and thus at increased risk of Diabetes Mellitus and atherosclerotic cardiovascular disease. In South Asian countries, the prevalence of type II Diabetes Mellitus and cardiovascular disease is very high ranging from 14 to 33% possibly due to type of food intake with high sodium content.(13)

The association between the TyG index and ASCVD events is reported in many studies, identifying it as a useful predictor of atherosclerosis. A study in Korea showed that the TyG index was an independent predictive marker of coronary atherosclerosis progression. Second, a case-control study from Taipei reported that the TyG index can predict the intima-media thicknesses of common carotid arteries. This relationship was studied in different populations, such as individuals with hypertension and diabetes (1) this index show high sensitivity and specificity for detecting insulin resistance. It also shows a link between with cardiovascular disease risk. (14)

TyG index showed a close connection with Diabetes Mellitus and cardiovascular disease so it can be used as an independent predictive marker of atherosclerosis. a higher TyG index is associated with a higher risk of CVD and mortality in the young population.(2) IR causes arteriosclerosis due to chronic hyperinsulinemia. Chronic hyperinsulinemia increases the synthesis of very low-density lipoprotein cholesterol, proliferation of vascular smooth muscle cells, transport of LDL cholesterol into arterial smooth muscle cells, and activation of inflammation genes.24 In addition, IR stimulates the progression of CAD by disrupting glucose metabolism, weakening systemic lipid metabolism, and causing endothelial dysfunction.

In this study, a significant correlation was found for high triglyceride glucose index with three vessels disease of the heart i.e. atherosclerosis. It leads to adverse cardiovascular outcomes like myocardial infarction and other ischemic diseases. Percutaneous intervention of the heart that showed no atherosclerosis, was associated with normal triglyceride glucose index i.e. <9.04. The triglyceride glucose index is strongly associated with Diabetes Mellitus. Fasting blood glucose and HbA1c were used for evaluation. The high triglyceride glucose index is due to insulin resistance which leads to vascular inflammation and stiffness at the tunica media. It results in vascular endothelial functional disorder. (15)

A U.S. study reported a high prevalence of Diabetes Mellitus in South Asians, with or without obesity. Diabetic patients were less likely to achieve adequate glycemic control.

The results of the study show a high TyG index in males as compared to females in the age group of 50 years and above. However, in females, the TyG index shows a similar pattern with higher TyG in the older population i.e. more than 50 years of age. A study by Liu Y et.al also demonstrated similar statistics of gender distribution in China population. In females heart disease onset occurs much later than men, due to vascular protective action of estrogen, which prevents atherosclerosis. Estrogen binding to receptors (ERs) promote vasodilation and reduce the response of blood vessels to the progression of atherosclerosis. (16)

The association of high TyG index with serum fasting triglycerides and cholesterol is also significant showing a p-value of <0.001, while LDL and HDL were non-significant.

From this study, we can surely determine that triglyceride glucose index is a reliable marker of insulin resistance which is responsible for poor action of insulin at target organs. New studies are proposing that insulin resistance can lead to vascular endothelial dysfunction and calcification leading to atherosclerosis. Atherosclerosis progresses to adverse cardiovascular outcomes and increases morbidity and mortality.

We and others have recently started to assess the relationship of a polygenic predictor of ASCVD in South Asian individuals and establish larger reference populations that may identify common or rare genetic factors distinct to South Asian individuals.

## Limitations

This study has the limitations of small sample size. The diagnostic modality used for atherosclerosis is not a single one i.e. by percutaneous intervention (PCI) and left heart catheterization (LHC).

## Data Availability

All data produced in the present study are available upon reasonable request to the authors

https://submit.medrxiv.org/submission/queue?queueName=submission_in_progress

